# Ancestry-Calibrated Polygenic Risk Scores Predict PTSD Trajectories in Recent Trauma Survivors and Interact with Neighborhood Resources

**DOI:** 10.64898/2026.07.17.26358149

**Authors:** E. Kate Webb, Aarti Jajoo, Vijetha Balakundi, Mohammad S.E. Sendi, Karestan C. Koenen, Sarah D. Linnstaedt, Stacey L. House, Xinming An, Jennifer S. Stevens, Thomas C. Neylan, Gari D. Clifford, Tanja Jovanovic, Laura T. Germine, Scott L. Rauch, John P. Haran, Alan B. Storrow, Christopher Lewandowski, Paul I. Musey, Phyllis L. Hendry, Sophia Sheikh, Christopher W. Jones, Brittany E. Punches, Lauren A. Hudak, Jose L. Pascual, Mark J. Seamon, Elizabeth M. Datner, Claire Pearson, Roland C. Merchant, Robert M. Domeier, Niels K. Rathlev, Brian J. O’Neil, Paulina Sergot, Leon D. Sanchez, Steven E. Bruce, Steven E. Harte, Ronald C. Kessler, Samuel A. McLean, Kerry J. Ressler, Nikolaos P. Daskalakis, Nathaniel G. Harnett

**Affiliations:** Division of Depression and Anxiety Disorders, McLean Hospital, Belmont, MA, 02478, USA; Department of Psychiatry, Harvard Medical School, Boston, MA, 02115, USA; Duke University School of Medicine, Department of Psychiatry, Durham, NC, USA; Stanley Center for Psychiatric Research, Broad Institute of MIT and Harvard, Cambridge, MA, 02142, USA; Tri-Institutional Center for Translational Research in Neuroimaging and Data Science (TReNDS), Georgia State University, Georgia Institute of Technology, Emory University, Atlanta, GA, 30302, USA; Department of Epidemiology, Harvard T.H. Chan School of Public Health, Harvard University, Boston, MA, 02115, USA; Institute for Trauma Recovery, Department of Anesthesiology, University of North Carolina at Chapel Hill, Chapel Hill, NC, 27559, USA; Department of Emergency Medicine, Washington University School of Medicine, St. Louis, MO, 63110, USA; Department of Psychiatry and Behavioral Sciences, Emory University School of Medicine, Atlanta, GA, 30329, USA; Departments of Psychiatry and Neurology, University of California, San Francisco, San Francisco, CA, 94143, USA; Department of Biomedical Informatics, Emory University School of Medicine, Atlanta, GA, 30332, USA; Department of Biomedical Engineering, Georgia Institute of Technology and Emory University, Atlanta, GA, 30332, USA; Department of Psychiatry and Behavioral Neurosciences, Wayne State University, Detroit, MI, 48202, USA; Institute for Technology in Psychiatry, McLean Hospital, Belmont, MA, 02478, USA; The Many Brains Project, Belmont, MA, 02478, USA; Department of Emergency Medicine, University of Massachusetts Chan Medical School, Worcester, MA, 01655, USA; Department of Emergency Medicine, Vanderbilt University Medical Center, Nashville, TN, 37232, USA; Department of Emergency Medicine, Henry Ford Health System, Detroit, MI, 48202, USA; Department of Emergency Medicine, Indiana University School of Medicine, Indianapolis, IN, 46202, USA; Department of Emergency Medicine, University of Florida College of Medicine -Jacksonville, Jacksonville, FL, 32209, USA; Department of Emergency Medicine, Cooper Medical School of Rowan University, Camden, NJ, 08103, USA; Department of Emergency Medicine, Ohio State University College of Medicine, Columbus, OH, 43210, USA; Ohio State University College of Nursing, Columbus, OH, 43210, USA; Department of Emergency Medicine, Emory University School of Medicine, Atlanta, GA, 30329, USA; Department of Surgery, Department of Neurosurgery, University of Pennsylvania, Philadelphia, PA, 19104, USA; Perelman School of Medicine, University of Pennsylvania, Philadelphia, PA, 19104, USA; Department of Surgery, Division of Traumatology, Surgical Critical Care and Emergency Surgery, University of Pennsylvania, Philadelphia, PA, 19104, USA; Department of Emergency Medicine, Jefferson Einstein Hospital, Jefferson Health, Philadelphia, PA, 19141, USA; Department of Emergency Medicine, Sidney Kimmel Medical College, Thomas Jefferson University, Philadelphia, PA, 19107, USA; Department of Emergency Medicine, Wayne State University, Ascension St. John Hospital, Detroit, MI, 48202, USA; Department of Emergency Medicine, Brigham and Women’s Hospital, Boston, MA, 02115, USA; Department of Emergency Medicine, Trinity Health-Ann Arbor, Ypsilanti, MI, 48197, USA; Department of Emergency Medicine, University of Massachusetts Medical School-Baystate, Springfield, MA, 01107, USA; Department of Emergency Medicine, Wayne State University, Detroit Receiving Hospital, Detroit, MI, 48202, USA; Department of Emergency Medicine, McGovern Medical School at UTHealth, Houston, TX, 77030, USA; Department of Emergency Medicine, Harvard Medical School, Boston, MA, 02115, USA; Department of Psychological Sciences, University of Missouri - St. Louis, St. Louis, MO, 63121, USA; Department of Anesthesiology, University of Michigan Medical School, Ann Arbor, MI, 48109, USA; Department of Internal Medicine-Rheumatology, University of Michigan Medical School, Ann Arbor, MI, 48109, USA; Department of Health Care Policy, Harvard Medical School, Boston, MA, 02115, USA; Department of Emergency Medicine, University of North Carolina at Chapel Hill, Chapel Hill, NC, 27559, USA; Department of Pharmacology, Physiology and Biophysics, Boston University Chobanian & Avedisian School of Medicine, Boston, MA, 02118, USA; Department of Medicine (Biomedical Genetics), Boston University Chobanian & Avedisian School of Medicine, MA, 02118, USA

**Keywords:** posttraumatic stress disorder, polygenic risk scores, neighborhood disadvantage, greenspace

## Abstract

**Objective:** Polygenic risk scores (PRS) for posttraumatic stress disorder (PTSD) often account for a low amount of variance. Ancestry-related differences in PRS scale and variance limit cross-group comparisons. This methodological challenge further complicates gene-by-environment (GxE) analyses, given that socioenvironmental exposures are inequitably distributed across ethnoracial groups. We constructed an ancestry-calibrated polygenic risk score (AC-PRS) for PTSD in the largest longitudinal study of trauma survivors to date and investigated GxE interactions.

**Method:** Recent trauma survivors (N=1,801) provided a blood specimen for genotyping. Six PTSD trajectories were previously identified from PTSD Checklist for DSM-5 (PCL-5) scores at 2-weeks, 8-weeks, 3-months, and 6-months post-trauma. Greenspace (normalized difference vegetation index [NDVI) and socioeconomic disadvantage (area deprivation index [ADI]) were derived from residential addresses. Logistic regressions examined interactions between newly developed AC-PRS and neighborhood factors on trajectories after adjusting for sociodemographic and trauma-related covariates. Secondary linear models considered GxE interactions on 6-month PCL-5 scores.

**Results:** AC-PRS performed well across ethnoracial groups, explaining significant variability in PTSD trajectories (R^2^=.053). ADI moderated the association between AC-PRS and the likelihood of assignment in a high nonremitting trajectory of PTSD symptoms and severity of symptoms at 6-months (*ps* < .05). There were no NDVI x AC-PRS interactions in any models.

**Conclusions:** AC-PRS captures genetic risk for PTSD in admixed trauma survivors, demonstrating good discrimination between nonremitting and resilient courses of PTSD. However, neighborhood disadvantage may modify utility of PRS for PTSD, warranting careful consideration when applying these scores across contexts.

## Introduction

Of the 40 million individuals who visit a US emergency department following a traumatic event each year, a subset will develop posttraumatic stress disorder (PTSD), a debilitating condition with significant psychological, financial, and social consequences^1^. Interventions administered in the acute aftermath of the trauma may help mitigate symptom development^2^; however, identifying those at risk for PTSD and who may benefit from early treatment is limited by both symptom heterogeneity and the complex interplay between biological predisposition, individual characteristics, and socioenvironmental factors^3^. Quantifying an individual’s genetic vulnerability to PTSD may be one approach to improve early identification of trauma survivors at-risk for a chronic and severe course of PTSD versus a resilient trajectory (i.e., low or no symptoms over time); however, no studies to date have examined the predictive utility of PRS-PTSD scores in a diverse, admixed sample of recent trauma survivors or investigated how genetic risk interacts with socioenvironmental factors on symptom trajectories.

Polygenic risk scores for PTSD (PRS-PTSD) aim to quantify individual genetic risk for the condition by aggregating across the small effects of many genetic variants (single-nucleotide polymorphisms [SNPS]) in the genome^4^. PRS-PTSD are derived by summing the individual’s SNP-level genotype by corresponding effect sizes from genome-wide association study (GWAS) summary statistics, but the performance of the scores decays when there are significant genetic differences between the GWASs and the target sample^5^. To date, only two studies have examined whether PRS-PTSD predicts future symptoms following a civilian traumatic event (e.g., motor vehicle collision)^6,7^. Notably, because the majority of available GWASs have been conducted in European-ancestry samples, PRS-PTSD often underperforms in non-European-ancestry populations, as it has been observed for PRS of other traits. For example, prior work found PRS-PTSD explains approximately 5% of PTSD symptom severity and onset in European-ancestry participants but fails to significantly predict PTSD status in African-ancestry participants^5,6,8^.

Consequently, the majority of gene-by-environment (G x E) studies are also ancestrally and ethnoracially biased due to disproportionate recruitment of non-Hispanic White individuals of European ancestry in GWASs^9^. Careful examination of G x E interactions between PRS-PTSD and socioenvironmental factors in ethnoracially diverse samples is critical for two reasons. First, there are significant ethnoracial inequities in trauma exposure and PTSD, with non-Hispanic Black trauma survivors showing significantly higher prevalence rates of PTSD compared to non-Hispanic White trauma survivors^10,11^. Second, exposure to socioenvironmental factors is not randomly distributed across people or places. Non-Hispanic Black and Hispanic individuals are disproportionately exposed to socioenvironmental risk factors, which may contribute to cumulative risk for PTSD^12–15^. Therefore, understanding how genetic profiles and socioenvironmental factors interact in trauma survivors of various ethnoracial backgrounds is critical for advancing health equity and requires PRS-PTSD with strong cross-ancestry comparability.

Recent advancements in PRS construction methods have sought to address cross-ancestry portability within the confines of available GWAS data^16–18^. For example, multi-ancestry methods such as PRS-CSx^17^, which integrate GWAS summary statistics from multiple populations, have demonstrated improved prediction for several continuous and highly heritable traits, including height, weight, and schizophrenia^19^. However, the extent to which these approaches generalize to PTSD, a disorder characterized by modest SNP heritability and strong environmental interactions, remains unclear. Here, we first present a new ancestry-calibrated PRS (AC-PRS) framework that harmonizes ancestry-matched PRS (PRS for admixed African (AA) samples derived from AA GWAS and PRS for European (EUR) samples derived from European GWAS) across populations to improve the cross-ancestry comparability of genetic risk.

The relative impact of socioenvironmental factors, including the socioeconomic and physical characteristics of a neighborhood, on the response to traumatic stress may depend, in part, on genetic variation^6,20^. For example, individuals in socioeconomically disadvantaged neighborhoods who also have a greater genetic risk for PTSD may face a “double jeopardy” and experience more severe and chronic outcomes than “single risk” individuals^6^. One study demonstrated that, for non-Hispanic White trauma survivors who were considered at high genetic risk for PTSD (but not “low risk” or “mid-risk”), living in a disadvantaged neighborhood was associated with an increased likelihood of developing PTSD, compared to those living in an advantaged neighborhood^6^. Prior G x E studies have also been limited by reliance on case-control designs, typically comparing individuals with chronic PTSD to healthy controls, which cannot establish how these interactions contribute to the *development* of PTSD. In other words, G x E studies rarely investigate how interactions with the broader context in which individuals live and trauma occurs influence risk for PTSD.

Prior work suggests that socioenvironmental risk factors may influence whether genetic risk is expressed following trauma, yet far less is known about the neighborhood characteristics that may instead promote resilience. Emerging work reveals that exposure to greenspace may facilitate recovery after trauma^12^; however, no study to date has examined G x E interactions in PTSD with residential greenspace. Considering how potential socioenvironmental protective factors may moderate genetic associations with PTSD may help identify factors that could be targeted to reduce the population burden of the condition.

The present study combined genetic and geospatial analyses of data collected as part of a large, longitudinal, multi-site study of recent trauma survivors (the AURORA Study)^21^ to investigate G x E interactions on PTSD trajectories. We first constructed the AC-PRS and established its predictive utility. Next, we investigated whether residential greenspace and neighborhood disadvantage moderated the association between AC-PRS and the likelihood of assignment in one of 6 previously identified PTSD trajectories^22^. Based on previous work, we hypothesized that individuals with greater genetic risk who resided in more disadvantaged neighborhoods would have a greater likelihood of assignment in a nonremitting PTSD trajectory, whereas individuals with lower genetic risk and more exposure to greenspace would have an increased likelihood of a resilient trajectory. We also considered the ethnoracial differences in socioenvironmental exposures and how this affected the predictive utility of AC-PRS, particularly among non-Hispanic Black and Hispanic trauma survivors.

## Methods

### Participants

Individuals were recruited from participating emergency departments (EDs) within 72 hours of a traumatic event as part of the AURORA Study^21^ (NIMH U01 MH110925; **Figure 1A**). Participants qualified for the study if they were between 18 and 65 years old and experienced a motor vehicle collision, physical assault, sexual assault, fall from greater than 10 feet, or mass casualty incident. Other trauma exposures were considered if the individual endorsed that they experienced, witnessed, or learned of actual or threatened serious injury, sexual violence, or death, and if the research team agreed that the event was a plausible Criterion A qualifying event. Complete details of the larger AURORA study are reported elsewhere^21,23,24^. Procedures were approved by each site’s institutional review board and all participants provided written informed consent and were financially compensated.

**Figure 1.**
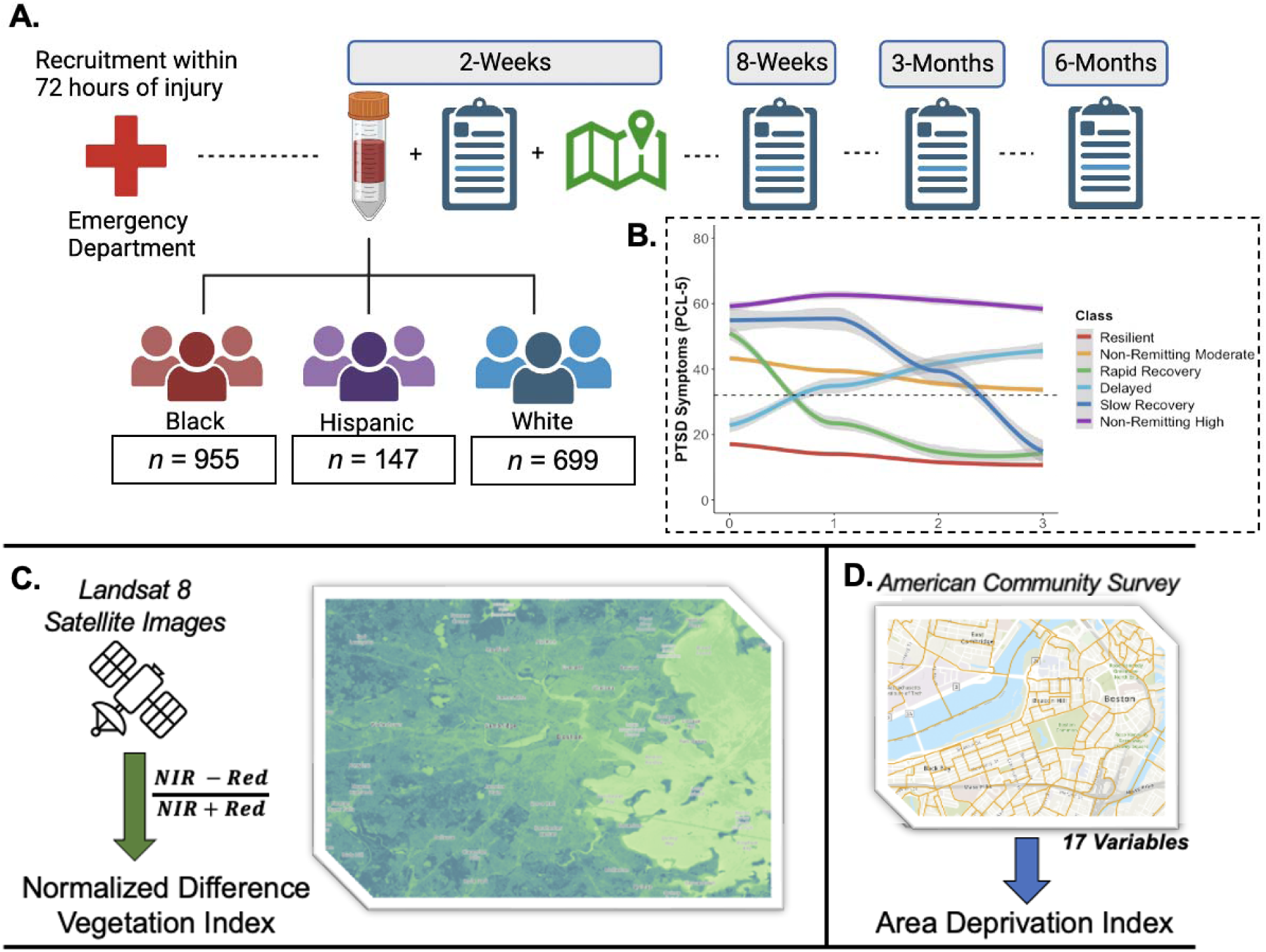
Study overview. **[A]** Trauma survivors were enrolled in a longitudinal cohort study in the Emergency Department and provided a blood specimen used for genotyping. Participants completed follow-up PTSD assessments at 2-weeks, 8-weeks, 3-months, and 6-months post-trauma, which were used to identify **[B]** six PTSD trajectories. **[C]** Residential greenspace was derived from high-resolution satellite imagery and quantified using the Normalized Difference Vegetation Index (NDVI). **[D]** Participants’ home addresses were assigned to a census block group and matched to the corresponding Area Deprivation Index (ADI), a 17-factor composite ranking of neighborhood socioeconomic disadvantage.

### Procedure

#### Genotyping

Participants provided a blood specimen at the time of enrollment or during the first follow-up visit using PAXgene DNA tubes. Then, genotyping was performed using the Infinium Multi-Ethnic Global Array (MEGA, Illumina). Complete details about genotyping processing can be found in **Supplemental Material**.

#### Ancestral Groups

We calculated ancestry proportions by projecting genotype data onto principal components derived from known continental-level ancestry markers (see **Supplemental Material**). Participants were grouped by ancestry proportions based on EUR > 0.90 and AA > 0.60: European ancestry (EUR; *n* = 913), admixed African ancestry (AA; *n* = 1,275), and Other (*n* = 548). The 548 individuals in the Other category were further subdivided into: 239 Admixed European (AE; EUR ancestry between 0.60 and 0.90), 180 with intermediate African ancestry (AFR between 0.40 and 0.60, labeled AA04–AA06), and 129 with East Asian, Central/South Asian, or American ancestries.

### Measures

#### Demographics

Participants completed a comprehensive demographics form in the ED, which included sex assigned at birth, age, education, ethnoracial group, and their residential address.

Ethnoracial Group was a combined self-reported race and ethnicity variable that included non-Hispanic White, non-Hispanic Black, Hispanic, and Other categories. The “Other” ethnoracial group category encompassed multiple responses, including non-Hispanic Asian, American Indian, Hawaiian or Pacific Islander, and multiracial. Due to the small sample size across various groups, extrapolating findings to a specific population was not possible; therefore, individuals in the Other category were excluded. At the 2-week visit, participants reported annual household income, which was converted into a semi-continuous variable in $20,000– $25,000 increments per unit increase.

#### PTSD Trajectories

The PTSD Checklist for DSM-5 (PCL-5)^25^ was administered to evaluate the presence and severity of PTSD symptoms at 2-weeks, 8-weeks, 3-months, and 6-months after trauma exposure. The 2-week survey queried symptoms experienced in the past two weeks (i.e., since the event). The other assessments referenced the past 30 days. Complete details on the latent-class mixed-effect modeling to derive PTSD trajectories are described in ^22^. Briefly, participants who completed the PCL-5 for at least 2 of the 4 time points were included, and 1 to 7 classes were tested. A 6-class solution with linear and quadratic terms provided the best fit to the data and included resilient, nonremitting high, nonremitting moderate, delayed, rapid recovery, and slow recovery classes^22^ (**Figure 1B**).

#### Neighborhood Factors

Complete details on deriving the neighborhood factors can be found in the **Supplemental Methods**. As previously conducted^22^, greenspace was quantified using the normalized difference vegetation index (NDVI) derived from high-resolution satellite imagery (Landsat-8 courtesy of the U.S. Geological Survey, extracted using Google Earth Engine^26,27^; **Figure 1C**). Mean NDVI values (range 0–1) within a 100-meter buffer around each residential address were extracted, with higher scores indicative of more greenspace.

The Area Deprivation Index (ADI; Version 3.1, 2019, downloaded from https://www.neighborhoodatlas.medicine.wisc.edu/))^28–30^ was used to quantify neighborhood socioeconomic disadvantage. The ranking (range 1–100) reflects each census block group’s socioeconomic position relative to all U.S. block grops, with a national ADI rank of 1 indicating the most advantaged neighborhood in the country.

#### Ancestry-Calibrated Polygenic Risk Scores (PRS)

We utilized EUR- and AA-specific PTSD Freeze 3 GWAS summary statistics^31^. It should be noted that since AURORA was one of the cohorts used in Freeze 3, we used the GWAS version provided by the Nievergelt Lab that excluded the AURORA cohort from the calculations. We masked the GWAS for chr3p21, chr6p21, and chr17q21.31 regions. Filtered SNPs shared between the EUR and AA genotype groups were used to calculate polygenic risk scores (PRS) using PRScs^32^. We used all default parameters of PRScs but for phi. *phi* is a global shrinkage parameter. As recommended, we used multiple phi values to generate PRS scores: e-6, e-4,.01 and 1. phi 0.01 for EUR GWAS and phi 0.1 for AA GWAS were chosen for downstream analysis based on PRS performance in distinguishing the resilient vs non-remitting moderate and non-remitting high groups, which were defined using temporal PCL-5 scores as described above.

To enable cross-ancestry comparison of PRS, we performed residualization and standardization in two strata: (i) EUR and (ii) a pooled AA, AE, and AA04AA06. Within each stratum, PRS were residualized on the first five ancestry PCs (PC1–PC5), and the resulting residuals were z-scored using the stratum-specific mean and standard deviation. Individuals from other ancestries (*n* = 129) were set to *NA* and excluded from downstream analyses.

#### Stress and Trauma-Related Assessments

In the ED, a physician recorded an injury severity score (ISS) quantifying the overall severity of all injuries (range: 0–75). A modified Childhood Trauma Questionnaire (mCTQ) evaluated childhood maltreatment at the 2-week visit^33^. Endorsements of each item were summed to create a total score (range: 0–44). Participants completed the Life Events Checklist for DSM-5 (LEC-5) at the 8-week visit^34^. The LEC-5 evaluated lifetime exposure to 17 different stressful/traumatic events. Participants were asked to indicate whether the event had happened to them, they had witnessed the event, or if they had learned of its occurrence. A total score was created by summing all responses (range: 0–68).

The 10-item Connor-Davidson Resilience Scale was administered at the 2-week visit to quantify perceived individual resources related to stress resiliency. Participants rated how accurately each of the statements (e.g., “Coping with stress can strengthen me”) described them on a scale of 0 (not true at all) to 4 (true nearly all the time; range: 0–40).

## Statistical Analysis

All analyses were completed in R (R version 4.1.2). The predictive utility of the AC-PRS was first evaluated by examining the proportion of variance (Nagelkerke’s R²) explained in PTSD trajectories (see **Supplemental Material**). Specifically, we compared the resilient trajectory with the non-remitting moderate and high trajectories, both individually and combined into a single non-remitting group. These groups were selected to maximize contrast between individuals characterized by persistently clinically elevated symptoms and those with low/no symptoms across time.

Participants were included in the primary G x E analyses if they had an AC-PRS, a PTSD trajectory assignment, complete self-report data, and were successfully geocoded (Study Flow Chart in **eFigure 1**). Bivariate associations between AC-PRS, PTSD symptoms, and neighborhood factors were compared using Pearson’s correlation tests. Ethnoracial differences in variables were probed using one-way ANOVAs with Tukey post-hoc tests.

We then examined the performance of AC-PRS in predicting PTSD trajectories by ethnoracial group. A multinomial logistic regression (package *nnet*) evaluated an Ethnoracial Group x AC-PRS interaction term on PTSD trajectory assignment after adjusting for sex (0 = male; 1 = female), age, education, income, ISS, lifetime trauma, childhood maltreatment, perceived individual resources, ADI, NDVI, and the top two PCs. Ethnoracial Group was dummy-coded, and non-Hispanic White was set as the reference group. AC-PRS and the first two genetic PCs were *z*-scored, and all other continuous dependent variables were mean-centered across the full sample. Covariates were selected based on their independent predictive utility for PTSD trajectories^22^.

The primary analysis tested two multinomial logistic regressions evaluating interactions between NDVI x AC-PRS as well as ADI x AC-PRS on trajectories, adjusting for the aforementioned covariates. For all logistic regressions, we used Wald z tests to assess statistical significance at *p* < .05. Finally, to complement the trajectory-based findings, we conducted two general linear models (GLMs) to examine interactions between AC-PRS and neighborhood factors on 6-month PCL-5 total scores (*n* = 1,395 with available 6-month PCL-5). Interaction effects were further probed using simple slopes tests (*reghelper* package) and by considering the effect of the AC-PRS at -1 standard deviation (SD) below the mean, at the mean, and +1 SD above the mean of the socioenvironmental factor. For these interaction models, a threshold of *p* < .025 was used, with Bonferroni correction applied across the two interaction models.

## Results

### Sample characteristics

Sample characteristics of participants included in the primary G x E analyses (N = 1,801; 64.46% female; 38.81% non-Hispanic White, 8.16% Hispanic, 53.03% non-Hispanic Black) are presented in **Table 1**.

**Table 1.** Sample characteristics.

| *Variable* | *% (n) or Mean (SD)* |  |  |  |
| --- | --- | --- | --- | --- |
| *Ethnoracial Group* | *Non-Hispanic White*<br>*n = 699* | *Hispanic*<br>*n = 147* | *Non-Hispanic Black*<br>*n = 955* | *ANOVAS/ $\chi^2$*<br>*Tests* |
| Sex at birth |  |  |  |  |
| Female | 63.37% (443) | 53.06% (78) | 67.01% (640) | $\chi^2(2, N = 1801) = 11.42, p = .003$ |
| Mechanism of Injury |  |  |  |  |
| Motor vehicle collision | 68.53% (479) | 74.90% (110) | 78.43% (749) | $\chi^2(8, N = 1801) = 86.33, p < .001$ |
| Physical Assault | 5.29% (37) | 10.88% (16) | 11.41% (109) |  |
| Fall | 11.73% (82) | 5.44% (8) | 4.19% (40) |  |
| Animal related | 3.86% (27) | 2.72% (4) | 1.36% (13) |  |
| Other<sup>+</sup> | 10.59% (74) | 6.12% (9) | 4.61% (44) |  |
| Age (years) | 39.36 (14.56) | 35.35 (12.33) | 36.09 (12.80) | $F(2, 1798) = 13.54, p < .001$ |
| Education (highest grade) | 15.90 (2.46) | 14.80 (3.02) | 14.57 (2.29) | $F(2, 1798) = 61.46, p < .001$ |
| Income | 3.22 (1.73) | 2.21 (1.33) | 2.04 (1.23) | $F(2, 1798) = 137.70, p < .001$ |
| Area Deprivation Index | 49.97 (24.91) | 61.32 (25.85) | 76.18 (24.15) | $F(2, 1798) = 231, p < .001$ |
| Normalized Difference Vegetation Index | 0.49 (0.14) | 0.39 (0.13) | 0.44 (0.12) | $F(2, 1798) = 56.70, p < .001$ |
| Percieved individual resources (CD-RISC) | 22.53 (7.47) | 21.72 (8.44) | 22.94 (8.37) | $F(2, 1798) = 1.68, p = .186$ |
| 2-Week PTSD Symptom Severity (PCL-5) | 30.05 (18.20) | 31.71 (20.07) | 31.29 (19.42) | $F(2, 1798) = 0.98, p = .374$ |
| 6-month PTSD symptom severity (PCL-5)* | 22.25 (18.47) | 25.92 (18.60) | 23.94 (19.00) | $F(2, 1392) = 2.31, p = .100$ |
| Lifetime trauma (LEC-5 total score) | 11.76 (9.68) | 11.01 (11.23) | 8.63 (8.63) | $F(2, 1798) = 23.72, p < .001$ |
| Childhood trauma (mCTQ) | 9.16 (9.60) | 11.19 (10.68) | 9.30 (9.93) | $F(2, 1798) = 2.68, p = .070$ |
| AC-PRS | -0.02 (1.01) | 0.03 (1.13) | -0.01 (0.97) | $F(2, 1798) = 0.12, p = .887$ |
| PTSD Trajectory | | | | $\chi^2(10, N = 1801) = 11.56, p = .315$ |
| Resilient | 51.35% (359) | 46.26% (68) | 51.10% (488) |  |
| Delayed | 5.15% (36) | 8.16% (12) | 3.77% (36) |  |
| Moderate Nonremitting | 29.18% (204) | 27.21% (40) | 27.54% (263) |  |
| High Nonremitting | 7.44% (52) | 8.16% (12) | 9.42% (90) |  |
| Rapid Recovery | 4.29% (30) | 7.49% (11) | 5.34% (51) |  |
| Slow Recovery | 2.58% (18) | 2.72% (4) | 2.83% (27) |  |

*Note:* + The other category included: non-motorized collisions, sexual assaults, incidents causing traumatic stress exposure to many people (e.g., plane crash), poisoning, burns, and other traumatic injuries. \* $n = 1395$ with 6-month PCL-5 scores. *Abbreviations:* **CD-RISC:** Connor-Davidson Resilience Scale; **CTQ:** modified-Childhood Trauma Questionnaire; **LEC:** Life Events Checklist for DSM-5; **AC-PRS:** ancestry-calibrated polygenic risk scores; **PCL-5:** PTSD Symptom Checklist for DSM-5.

### Ancestry stratification reveals the need for AC-PRS

We first inferred continental-level ancestry proportions for all participants to stratify individuals by genetic background and evaluate ancestry-dependent differences in PRS distributions (**eFigure 2A–B**). As described in the Methods, based on the distribution of inferred ancestry proportions, individuals were classified into four groups (i..e, EUR, AE, AA, and AFR), while remaining individuals were excluded from downstream analyses (**eFigure 2C**, Methods).

We constructed PRS using summary statistics from the latest ancestry-specific GWAS^31^, appropriately excluding AURORA participants. When PRS were computed using ancestry-matched GWAS, we observed substantial differences in both the mean and variance of PRS distributions across ancestry groups (AA PRS mean = 0.06, standard deviation = 1.69; EUR PRS mean = -0.43, standard deviation = 0.16; **eFigure 2D**). These distributional differences persisted even when PRS were derived from the same GWAS and evaluated across different ancestry strata (**eFigure 2D**).

### AC-PRS significantly predicts PTSD trajectory assignment

To address the distributional differences, we adopted an ancestry-calibrated PRS (AC-PRS) framework. Specifically, PRS were first computed within each ancestry-matched GWAS using PRS-CS, followed by residualization with respect to ancestry principal components and subsequent standardization to enforce homoscedasticity across groups. Our PRS calibration substantially improved predictive performance across the full sample, as reflected by increased variance explained (resilient group vs. combined moderate/high non-remitting group; Null R^2^ = 0.036; uncalibrated PRS R^2^ = 0.045; AC-PRS R^2^ = 0.053; **Figure 2**).

**Figure 2.**
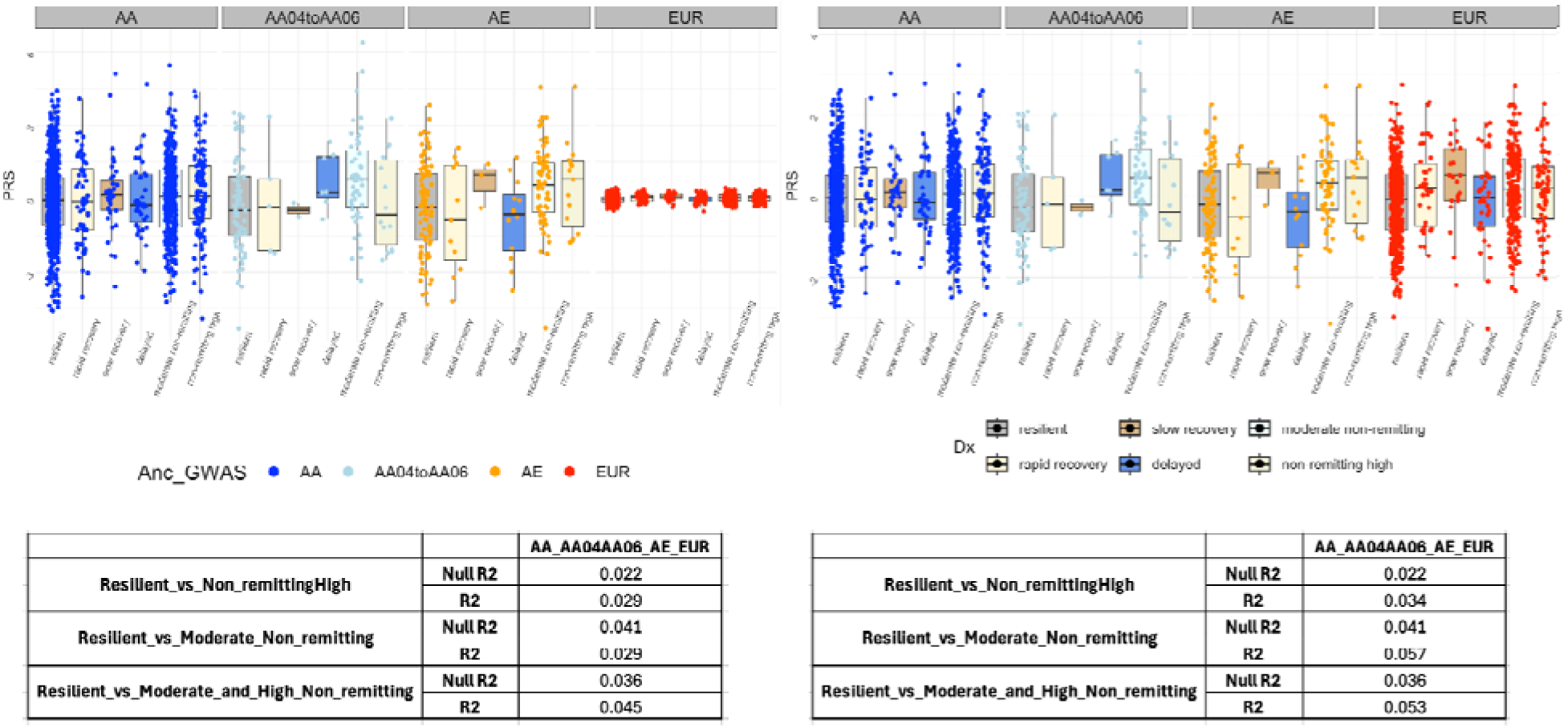
Ancestry calibration improves PRS comparability and predictive performance across ancestry groups. **[A].** PRS after residualization for ancestry principal components. **[B].** AC-PRS after standardization of the residualized PRS within ancestry groups, shown in **(A)**. Boxplot displays PRS distribution across recovery trajectory groups within each ancestry strata.

### AC-PRS is not associated with socioenvironmental inequities

There was no correlation between AC-PRS and ADI (*r*(1799) = 0.02*, p* = .464) or NDVI (*r*(1799) *=* 0.02, *p* = .418). Separate one-way ANOVAs revealed that both ADI and NDVI significantly varied by ethnoracial group (ADI: *F*(2, 1798) = 231, *p* < .001; NDVI: *F*(2, 1798) = 56.7, *p* < .001). Follow-up Tukey tests indicated non-Hispanic Black and Hispanic participants lived in more disadvantaged neighborhoods (**eFigure 3A**) compared to non-Hispanic White participants, and non-Hispanic Black participants resided in more disadvantaged neighborhoods compared to Hispanic participants (*ps* adjusted < .05). Further, non-Hispanic Black and Hispanic participants were exposed to significantly less greenspace (**eFigure 3B**) than non-Hispanic White participants, and non-Hispanic Black participants were exposed to less greenspace than Hispanic participants (*ps* adjusted < .05). A third ANOVA revealed there was no association between ethnoracial group on AC-PRS (**eFigure 3C**; *F*(2, 1798) = 0.12, *p* = .887).

**eTable 1** presents the results of a multinomial logistic regression examining AC-PRS on PTSD trajectories without any interaction terms. The model examining an AC-PRS x Ethnoracial group interaction on PTSD trajectories revealed that relationships between AC-PRS and trajectory assignment among Hispanic and non-Hispanic Black participants were comparable to those observed in non-Hispanic White participants (all non-Hispanic Black x AC-PRS and Hispanic x AC-PRS terms *ps* >L.05; **eTable 2**).

### Interactions between socioenvironmental factors and AC-PRS on PTSD trajectories

A multinomial logistic regression model examining an ADI x AC-PRS term revealed that ADI moderated the association between AC-PRS and the likelihood of assignment in the nonremitting-high trajectory versus a resilient trajectory (Wald *z* testL=L-2.29; *p* =L.022), even after adjusting for covariates (**Table 2**). Higher AC-PRS at lower levels of ADI was associated with greater likelihood of assignment to the non-remitting high compared to resilient trajectory (**eFigure 4**). There was no significant NDVI x AC-PRS interaction on trajectory assignment (**eTable 3**).

**Table 2.**
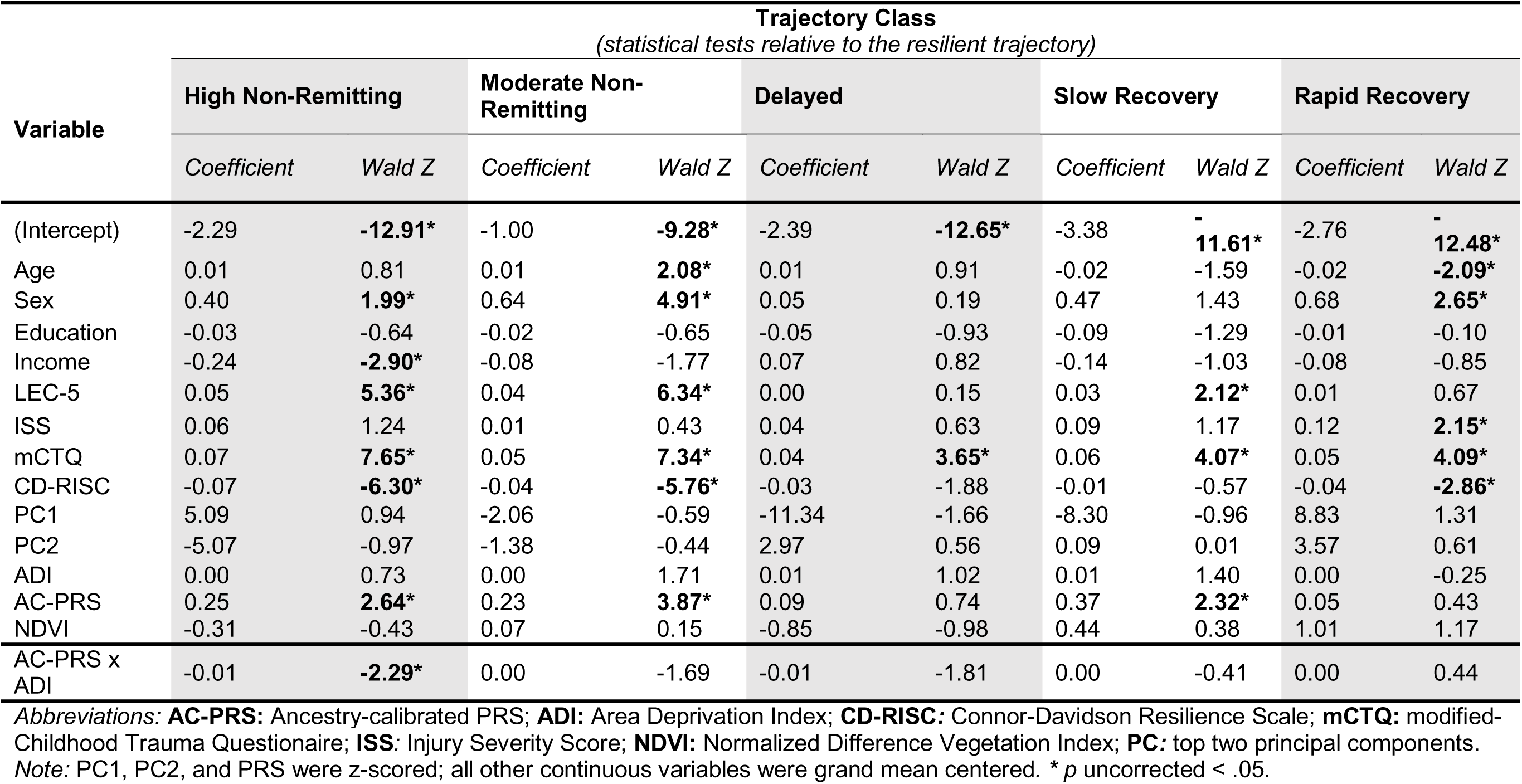
AC-PRS x ADI interaction on PTSD trajectory assignment.

### Interactions between socioenvironmental factors and AC-PRS on 6-month PTSD severity

Higher AC-PRS (*r*(1393) = 0.13*, p* < .001) and ADI (*r*(1393) = 0.11*, p* < .001) were associated with higher 6-month PCL-5 scores, whereas NDVI (*r*(1393) = -0.05*, p* = .047) was associated with lower 6-month PCL-5 scores. A GLM identified a significant AC-PRS x ADI interaction on 6-month PTSD symptoms after adjustment for covariates (interaction term: *t*(1380) = -2.62, *p* = .009; significant with Bonferroni correction). Consistent with the trajectory findings, results suggested that higher ADI attenuated the effect of AC-PRS on 6-month PTSD symptoms. The simple slopes tests (**Figure 3A**) demonstrated that higher AC-PRS was associated with more severe PTSD symptoms among trauma survivors living in neighborhoods with lower (b = 2.78, SE = 0.07, *p* < .001) and average ADI (b = 1.55, SE = 0.47, *p* = .001) but not higher ADI (b = 0.33, SE = 0.50, *p* = .617). There was no effect of AC-PRS x NDVI on 6-month symptoms after adjusting for covariates (interaction term: *t*(1380) = 0.53, *p* = .594; **Figure 3B**).

**Figure 3.**
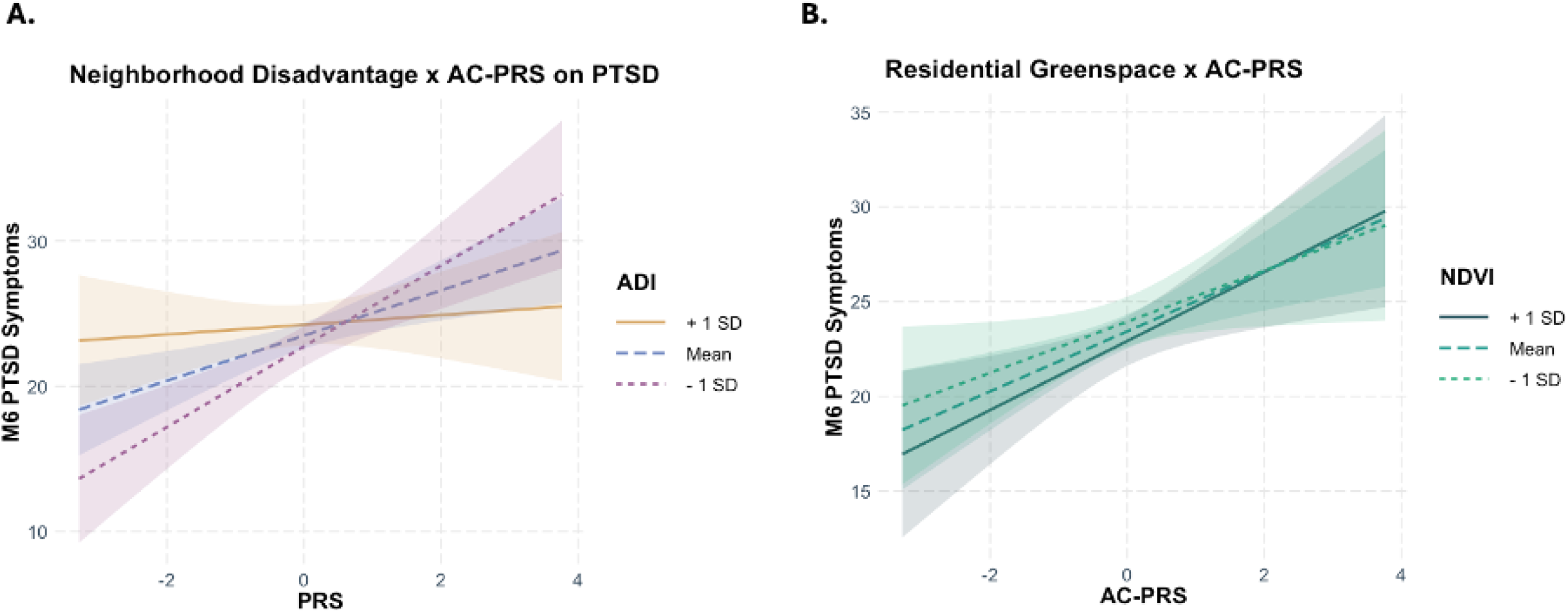
G x E interactions on 6-month PTSD symptom severity. **[A]** Higher AC-PRS was associated with more severe PTSD symptoms at lower (b = 2.78, SE = 0.07, *p* < .001) and average ADI (b = 1.55, SE = 0.47, *p* = .001) but not higher ADI (b = 0.33, SE = 0.50, *p* = .617). **[B]** There was no interaction between AC-PRS and residential greenspace on PTSD symptoms.

## Discussion

We report the first prospective cohort study to construct and evaluate the predictive utility of AC-PRS for PTSD among admixed recent trauma survivors. Our results suggest that AC-PRS may be a particularly useful method to calculate polygenic risk for PTSD that is less biased by socioenvironmental confounds and ancestral bias. The G x E interaction models revealed that neighborhood disadvantage modified the performance of AC-PRS in distinguishing between non-remitting high versus resilient trajectories. Further, higher AC-PRS was associated with 6-month PTSD symptoms only among individuals living in more advantaged neighborhoods. Our findings add to the existing literature examining acute predictors of PTSD trajectories, underscoring how neighborhood disadvantage may modify the relationship between genetic risk and symptom development and play a critical role in risk for PTSD.

Findings present a new AC-PRS framework to improve the cross-ancestry comparability of genetic risk for PTSD that may be adopted in future work. The highly polygenic architecture and modest SNP heritability of PTSD, together with differences in linkage disequilibrium structure and minor allele frequencies across populations, can introduce error when constructing PRS-PTSD^35^. Under these conditions, direct cross-ancestry portability of PRS can be limited. In contrast, ancestry-calibrated approaches, such as the one adopted herein, rely on ancestry-matched PRS to mitigate these sources of divergence by preserving within-ancestry genetic signal while harmonizing score distributions across populations. Findings demonstrate that by aligning PRS scaling rather than SNP-level effect estimation across ancestries, the AC-PRS improves comparability of genetic risk without assuming full transferability of genetic effects. In line with this interpretation, AC-PRS performed well across ethnoracial groups, explaining small but significant variability in PTSD trajectories (R^2^ = 0.053). In particular, higher AC-PRS was associated with increased likelihood of membership in the non-remitting high and moderate symptom trajectories, as well as the slow recovery trajectory, suggesting there is shared genetic risk underlying a protracted symptom course.

The AC-PRS and neighborhood disadvantage interaction results differ from those previously reported in Lobo et al. (2021), which found that for European-ancestry trauma survivors with high genetic risk, residing in a disadvantaged neighborhood increased the risk of PTSD compared to living in an advantaged neighborhood. In the present study, higher genetic risk was not associated with more severe PTSD symptoms at higher levels of neighborhood disadvantage. This finding may suggest that the influence of genetic risk on PTSD development is attenuated in highly disadvantaged neighborhoods where socioenvironmental risk factors, including those highly correlated with socioeconomic disadvantage (e.g., community violence, pollutants), may be more salient^36^. This is reminiscent of a seminal study by Ouellet-Morin and colleagues (2008), in which 19-month-old twins’ cortisol reactivity in conditions of high familial adversity was explained by both shared and unique environmental factors, but not genetic factors^37^. However, our observed patterns may also reflect a ceiling effect, as higher AC-PRS was significantly associated with greater 6-month PTSD symptoms. We did not observe a significant interaction between AC-PRS and greenspace. However, our prior work in this cohort suggests greenspace may interact with individual resources on PTSD trajectories and is associated with increased neural reactivity to rewarding stimuli^22^. Thus, suggesting greenspace may confer resilience through mechanisms independent of genetic susceptibility^22^. Ultimately, while the results highlight the genetic influences on PTSD risk, they also underscore that certain socioenvironmental factors may moderate this risk, changing how AC-PRS performs, and should not be overlooked.

In this prospective study, we demonstrate that AC-PRS can predict elevated symptom trajectories over six months post-trauma among individuals recruited in the immediate aftermath of a traumatic event. PRS for PTSD may support screening and personalized treatment for PTSD and possibly lead to the development of specific interventions for individuals with higher genetic risk^38–40^. However, the benefit of PRS’ clinical use for PTSD is tempered by its modest predictive power and challenges related to diversity and equity. The limited accuracy of traditional PRS among individuals of non-European ancestry is attributed to the lack of ancestral diversity in the large-scale GWASs and explained by differences in neutral human evolution (i.e., history of human migration, which leads to population stratification bias)^41,42^. Ongoing efforts to enhance cross-ancestry portability, including frameworks such as the AC-PRS presented here, as well as broader recruitment strategies and inclusion of non-European-ancestry participants in GWASs, may advance PRS’ clinical utility^42^.

In the United States, there are striking differences in exposure to neighborhood disadvantage and greenspace, not only between regions but also between ethnoracial groups, such that non-Hispanic Black and Hispanic individuals are disproportionately exposed to structural inequities^13,43^. Previous work has reported on the challenges of comparing PRS performance in different populations (e.g., across ancestral groups, in developmental and adult cohorts) and separating genetic influences on phenotypes from the role of socioenvironmental factors^44–48^. Several studies have emphasized that socioenvironmental factors may be captured in genetic components, which ultimately decreases prediction accuracy and can artificially inflate genetic-phenotype associations^49,50^. Taken together, our findings demonstrate that using methods that calibrate across ancestry may enhance the reliability of these scores^47^ and that testing associations and interactions with environmental confounds is a necessary methodological step.

## Limitations

Though the present study had multiple strengths, including the relatively large and well-characterized sample, several limitations temper the generalizability of the study findings. The overarching goal of AC-PRS was to help address GWAS inequities by leveraging statistical approaches to improve PRS performance in multi-ancestry samples. The GWASs leveraged to derive the AC-PRS included far fewer individuals with Hispanic/Latino ancestry compared to individuals with European- or even African-ancestry. Future work is needed to expand both the African and the Hispanic/Latino GWAS to ensure accurate and well-powered AC-PRS for PTSD estimates. More broadly, there is a need to stratify the performance of AC-PRS for various disorders and traits by ethnoracial group to ensure increases in prediction accuracy are not driven by a specific group. Biologically informed PRS, for example those relying on ancestry-specific or multi-ancestry TWAS, might be less susceptible to ancestry-specific differences^51^.

The GWASs used to derive PRS for PTSD distinguished between PTSD cases (lifetime and current) and controls^31^, whereas this study examined whether scores could forecast future symptom severity in only trauma-exposed individuals. In the current study, the AC-PRS was trained on PTSD symptom trajectories rather than a single timepoint. While we consider this design a major strength of the current study, examining the differences between the genetic profiles in chronic or lifetime PTSD cases (in GWASs) versus the profiles of prediction of PTSD acutely post-trauma is an important area for future research.

Given that the majority (∼75%) of trauma survivors experienced a motor vehicle collision, interactions between trauma type, socioenvironmental factors, and AC-PRS were not considered. As the largest studies examining PRS-PTSD have been conducted in single trauma samples (e.g., 9/11 responders^8^, motor vehicle collisions^6^, combat exposure^5^), future research should consider whether the performance of PRS varies by trauma type. To date, there is no validated approach to constructing PRS for multi-dimensional indicators of posttraumatic dysfunction, such as the Research Domain Criteria framework domains (e.g., negative valence). While we speculate that there is probably an overlap between the G x E interactions for PTSD and these domains, additional work is required to address the breadth of socioenvironmental factors and posttraumatic outcomes.

## Conclusions

The present findings offer a novel, ancestry-calibrated approach for predicting PTSD development with PRS in admixed samples. In a sample of recent trauma survivors, AC-PRS demonstrated strengths in differentiating between more chronic courses of PTSD compared to a resilient trajectory; however, neighborhood disadvantage moderated this effect. By leveraging AC-PRS, we could evaluate interactions between genetic and socioenvironmental factors on PTSD while mitigating ancestral bias and potential confounds with ethnoracial group. When interpreting PRS for PTSD, researchers should use ancestry-calibrated methods, paying close attention to whether prediction accuracy varies by ethnoracial groups or socioenvironmental context. To truly advance risk prediction models in PTSD, unbiased recruitment, equitable method development, and consideration of the socioenvironmental context must be the foundations of genomic approaches.

## Data Availability

Data and/or research tools used in the preparation of this manuscript were obtained from the National Institute of Mental Health (NIMH) Data Archive (NDA). NDA is a collaborative informatics system created by the National Institutes of Health to provide a national resource to support and accelerate research in mental health. Dataset identifier(s): NIMH Data Archive Digital Object Identifier (DOI) 10.15154/fqbt-fk91. This manuscript reflects the views of the authors and may not reflect the opinions or views of the NIH or of the Submitters submitting original data to NDA.

## Acknowledgments

The investigators would like to thank the trauma survivors participating in the AURORA Study. Their time and effort during a challenging period of their lives make our efforts to improve recovery for future trauma survivors possible. The content is solely the responsibility of the authors and does not necessarily represent the official views of any of the funders. Data and/or research tools used in the preparation of this manuscript were obtained from the National Institute of Mental Health (NIMH) Data Archive (NDA). NDA is a collaborative informatics system created by the National Institutes of Health to provide a national resource to support and accelerate research in mental health. Dataset identifier(s): NIMH Data Archive Digital Object Identifier (DOI) 10.15154/fqbt-fk91. This manuscript reflects the views of the authors and may not reflect the opinions or views of the NIH or of the Submitters submitting original data to NDA. Dr. Webb and Dr. Harnett had full access to all the data in the study and take responsibility for the integrity of the data and the accuracy of the data analysis.

## Funding

The present research was supported by NIMH T32MH017119 (EKW), T32MH125786 (MSES), K01MH119603 (NGH), U01MH110925 (SAM), R01MH106595 (AJ, VB, KCK, KJR and NPD) and R01MH133268 (AJ, VB, KJR and NPD); the US Army MRMC W81XWH-22-2-0078 (AJ, VB, KJR and NPD), One Mind, and The Mayday Fund. EKW received support from a Harvard Data Science Initiative Postdoctoral Fellow Research Fund award and a Phyllis and Jerome Lyle Rappaport Mental Health Research Scholar award.

## Disclosures

- Dr. Sendi receives consulting fees from NIJI Corp.
- Dr. Koenen’s research has been supported by the Robert Wood Johnson Foundation, the Kaiser Family Foundation, the Harvard Center on the Developing Child, Stanley Center for Psychiatric Research at the Broad Institute of MIT and Harvard, the National Institutes of Health, One Mind, Anonymous LLC, and Cohen Veterans Bioscience. She has been a paid consultant in the last three years for the US Department of Justice. She has had paid speaking engagements in the last three years with the American Psychological Association, European Central Bank. Sigmund Freud University – Milan, Cambridge Health Alliance, and Coverys. She receives royalties from Guilford Press and Oxford University Press.
- Dr. Neylan has received research support from NIH, VA, and Rainwater Charitable Foundation, and consulting income from Jazz Pharmaceuticals.
- In the last three years, Dr. Clifford has received research funding from the NSF, NIH, and LifeBell AI, and unrestricted donations from AliveCor Inc, Amazon Research, the Center for Discovery, the Gates Foundation, Google, the Gordon and Betty Moore Foundation, MathWorks, Microsoft Research, Nextsense Inc, One Mind Foundation, and the Rett Research Foundation. Dr Clifford has financial interest in AliveCor Inc and Nextsense Inc. He also is the CTO of MindChild Medical with significant stock. These relationships are unconnected to the current work.
- Dr. Jovanovic receives support from the National Institute of Mental Health, R01 MH129495.
- Dr. Germine receives funding from the National Institute of Mental Health (R01 MH121617) and is on the board of the Many Brains Project. Her family also has equity in Intelerad Medical Systems, Inc.
- Dr. Rauch reported serving as secretary of the Society of Biological Psychiatry; serving as a board member of Community Psychiatry and Mindpath Health; serving as a board member of National Association of Behavioral Healthcare; serving as secretary and a board member for the Anxiety and Depression Association of America; serving as a board member of the National Network of Depression Centers; receiving royalties from Oxford University Press, American Psychiatric Publishing Inc, and Springer Publishing; and receiving personal fees from the Society of Biological Psychiatry, Community Psychiatry and Mindpath Health, and National Association of Behavioral Healthcare outside the submitted work.
- Dr. Jones has no competing interests related to this work, though he has been an investigator on studies funded by AstraZeneca, Vapotherm, Abbott, and Ophirex.
- Dr. Datner serves as a Medical Advisor and on the Board of Directors for Cayaba Care.
- Dr. Harte has no competing interests related to this work, though in the last three years he has received research funding from Aptinyx and Arbor Medical Innovations, and consulting payments from Memorial Sloan Kettering Cancer Center, Indiana University, The Ohio State University, and Dana Farber Cancer Institute.
- In the past 3 years, Dr. Kessler was a consultant for Cambridge Health Alliance, Canandaigua VA Medical Center, Holmusk, Partners Healthcare, Inc., RallyPoint Networks, Inc., and Sage Therapeutics. He has stock options in Cerebral Inc., Mirah, PYM, and Roga Sciences.
- Dr. McLean served as a consultant for Walter Reed Army Institute for Research and for Arbor Medical Innovations.
- Dr. Ressler has performed scientific consultation for Bioxcel, Bionomics, Acer, and Jazz Pharma; serves on Scientific Advisory Boards for Sage, Boehringer Ingelheim, Senseye, and the Brain Research Foundation, and he has received sponsored research support from Alto Neuroscience.
- In the past 2 years, NPD has served on the Scientific Advisory Board of Circular Genomics, Jimini Health and Polaris Genomics, from which he has received consulting income and equity compensation, and received research support from Johnson & Johnson for unrelated work.

## Notes

### Competing Interest Statement

Disclosures:
Dr. Sendi receives consulting fees from NIJI Corp.
Dr. Koenen has been supported by the Robert Wood Johnson Foundation, the Kaiser Family Foundation, the Harvard Center on the Developing Child, Stanley Center for Psychiatric Research at the Broad Institute of MIT and Harvard, the National Institutes of Health, One Mind, Anonymous LLC, and Cohen Veterans Bioscience. She has been a paid consultant in the last three years for the US Department of Justice. She has had paid speaking engagements in the last three years with the American Psychological Association, European Central Bank. Sigmund Freud University, Milan, Cambridge Health Alliance, and Coverys. She receives royalties from Guilford Press and Oxford University Press.
Dr. Neylan has received research support from NIH, VA, and Rainwater Charitable Foundation, and consulting income from Jazz Pharmaceuticals.
In the last three years, Dr. Clifford has received research funding from the NSF, NIH, and LifeBell AI, and unrestricted donations from AliveCor Inc, Amazon Research, the Center for Discovery, the Gates Foundation, Google, the Gordon and Betty Moore Foundation, MathWorks, Microsoft Research, Nextsense Inc, One Mind Foundation, and the Rett Research Foundation. Dr Clifford has financial interest in AliveCor Inc and Nextsense Inc. He also is the CTO of MindChild Medical with significant stock. These relationships are unconnected to the current work.
Dr. Jovanovic receives support from the National Institute of Mental Health, R01 MH129495.
Dr. Germine receives funding from the National Institute of Mental Health (R01 MH121617) and is on the board of the Many Brains Project. Her family also has equity in Intelerad Medical Systems, Inc. Dr. Rauch reported serving as secretary of the Society of Biological Psychiatry; serving as a board member of Community Psychiatry and Mindpath Health; serving as a board member of National Association of Behavioral Healthcare; serving as secretary and a board member for the Anxiety and Depression Association of America; serving as a board member of the National Network of Depression Centers; receiving royalties from Oxford University Press, American Psychiatric Publishing Inc, and Springer Publishing; and receiving personal fees from the Society of Biological Psychiatry, Community Psychiatry and Mindpath Health, and National Association of Behavioral Healthcare outside the submitted work.
Dr. Jones has no competing interests related to this work, though he has been an investigator on studies funded by AstraZeneca, Vapotherm, Abbott, and Ophirex.
Dr. Datner serves as a Medical Advisor and on the Board of Directors for Cayaba Care.
Dr. Harte has no competing interest related to this work, though in the last three years he has received research funding from Aptinyx and Arbor Medical Innovations, and consulting payments from Memorial Sloan Kettering Cancer Center, Indiana University, The Ohio State University, and Dana Farber Cancer Institute.
In the past 3 years, Dr. Kessler was a consultant for Cambridge Health Alliance, Canandaigua VA Medical Center, Holmusk, Partners Healthcare, Inc., RallyPoint Networks, Inc., and Sage Therapeutics. He has stock options in Cerebral Inc., Mirah, PYM, and Roga Sciences.
Dr. McLean served as a consultant for Walter Reed Army Institute for Research and for Arbor Medical Innovations.
Dr. Ressler has performed scientific consultation for Bioxcel, Bionomics, Acer, and Jazz Pharma; serves on Scientific Advisory Boards for Sage, Boehringer Ingelheim, Senseye, and the Brain Research Foundation, and has received sponsored research support from Alto Neuroscience.
In the past 2 years, NPD has served on the Scientific Advisory Board of Circular Genomics, Jimini Health, and Polaris Genomics, from which he has received consulting income and equity compensation, and received research support from Johnson & Johnson for unrelated work.

### Author Declarations

Written informed consent was obtained from all participants as approved by each sites institutional review board, with review completed by the overall study site, the Institutional Review Board at the University of North Carolina at Chapel Hill.

