## Supplemental Material for "Ancestry-Calibrated Polygenic Risk Scores Predict PTSD Trajectories in Recent Trauma Survivors and Interact with Neighborhood Resources"

**eMethods.**

**eTable 1.** Effect of AC-PRS on PTSD trajectories.

**eTable 2.** AC-PRS x Ethnoracial Group interaction on PTSD trajectories.

**eTable 3.** AC-PRS x NDVI interaction on PTSD trajectories.

**eFigure 1.** Study flow chart.

**eFigure 2.** Genetic ancestry structure and ancestry-dependent behavior of PTSD PRS in the AURORA cohort.

**eFigure 3.** Ethnoracial differences in socioenvironmental factors and AC-PRS

**eFigure 4.** AC-PRS x ADI interaction on PTSD trajectories.

### eMethods

#### Calculating ancestry proportions.

We calculated ancestry proportions for each subject by projecting their genotype data onto principal components derived from known continental-level ancestry markers. For this, we used the global ancestry pipeline (https://github.com/nievergeltlab/global_ancestry), which applies SNPweights[1] to a set of 10,000 ancestry-informative markers. These markers were evaluated against a reference panel comprising 2,911 individuals from 71 diverse populations and six continental groups (k = 6)[2]. Participants (N = 2,736) were grouped by ancestry proportions: European ancestry (EUR > 0.90; n = 913), admixed African ancestry (AA > 0.60; n = 1,275), and Other (n = 548), based on ancestry proportions—EUR > 0.9 and AA > 0.6. The 548 individuals in the Other category were further subdivided into: 239 Admixed European (EUR ancestry between 0.6 and 0.9), 180 with intermediate African ancestry (AFR between 0.4 and 0.6, labeled AA04–AA06), and 129 with East Asian, Central/South Asian, or American ancestries. SNPs within each group with minor allele frequency (MAF > 0.1) are dropped. Genetic principal components were calculated using plink and pruned.

#### Genotype Imputation and Quality Control

Before pre-phasing and imputation, we first performed strand flipping according to our reference panel (1000G phase3 v5, GRch37/hg19) to improve imputation accuracy. Ambiguous SNPs (i.e., A/T or C/G SNPs) had already been removed in the pre-imputation QC step. For non-ambiguous SNPs, the alleles in our cohort were flipped if they appeared in the minus strand when compared with the reference panel (e.g., the alleles in our cohort are A/G, while they are T/C or C/T in the reference panel). Samples with genotype missingness >2% were excluded (PLINK --mind 0.02), and variants with missingness >2% were removed (--geno 0.02). Variants showing significant deviation from Hardy–Weinberg equilibrium ($p < 1 \times 10^{-6}$) were excluded. This resulted in genotype data of 2736 subjects.

The Michigan Imputation Server (https://imputationserver.sph.umich.edu) was used for phasing (via Eagle v2.4) and imputation (via Minimac4), using the HRC r1.1 reference panel. After imputation, we conducted an imputation quality assessment, which considered the imputation quality metric (dosage$R^2$: the squared Pearson correlation between the estimated allele dosages and the true experimental genotypes at the site and individual level). In the post-imputation filtering process, we retained only variants with imputation quality $R^2$ greater than 0.8, which were considered as "good quality" variants.

**Deriving Neighborhood Factors.**

As previously reported [3], residential greenspace was quantified using the normalized difference vegetation index (NDVI) derived from high-resolution (30m) multiband satellite imagery (Landsat-8 courtesy of the U.S. Geological Survey, extracted using Google Earth Engine [4,5]. Images acquired between May 2017 and September 2017 were used to capture the highest quality data and greenspace exposure prior to the study period. Images with less than 20% cloud cover were considered usable, and all images underwent preprocessing steps, including atmospheric correction and Top of Atmosphere reflectance conversion to remove effects from water vapor and sun position. NDVI was calculated with the following equation: NDVI = (near-infrared band – red band / near-infrared band + red band) [6].

The greenspace rasters and X-Y coordinates of the participants' home addresses were then entered into ArcGIS Pro Version 3.0.0 (ESRI, 2018). A 100m Euclidean buffer was created around each address. To avoid penalizing other natural infrastructure, bodies of water (negative NDVI) were set as null. Finally, mean NDVI values (range 0-1) within each buffer were extracted with zonal spatial analyses.

The Area Deprivation Index (ADI; Version 3.1, 2019, downloaded from https://www.neighborhoodatlas.medicine.wisc.edu/))[7–9] was used to quantify neighborhood socioeconomic disadvantage. Participants' home addresses were geocoded and matched to their respective census block group (the smallest publicly available geographical area). ADI is a weighted composite score of 17 items from the American Community Survey that represent domains of income, education, employment, and housing quality. The ranking (range 1 – 100) reflects each block group's socioeconomic position relative to all U.S. block groups, with a national ADI rank of 1 indicating the most advantaged neighborhood in the country.

***Statistical Analyses***

The predictive utility of the AC-PRS was first evaluated by examining the proportion of variance explained in groups identified by PTSD PCL-5 trajectories. We fitted logistic regression models, in which group status (e.g., resilient vs. non-remittent high) was the outcome and PRS was the predictor, adjusting for sex, age, and the first 10 genetic principal components (PCs). The performance was evaluated as the increase in Nagelkerke's $R^2$ between the full logistic regression model (group status ~ PRS + covariates) and the corresponding null model including only covariates (sex, age, and the first 10 genetic principal components).

**eTable 1.** Effect of AC-PRS on PTSD trajectory assignment

| Variable | Trajectory Class (statistical tests relative to the resilient trajectory) | | | | | | | | | |
|---|---|---|---|---|---|---|---|---|---|---|
| | **High Non-Remitting** | | **Moderate Non-Remitting** | | **Delayed** | | **Slow Recovery** | | **Rapid Recovery** | |
| | *Coefficient* | *Wald Z* | *Coefficient* | *Wald Z* | *Coefficient* | *Wald Z* | *Coefficient* | *Wald Z* | *Coefficient* | *Wald Z* |
| (Intercept) | -2.26 | -12.90* | -1.00 | -9.26* | -2.38 | -12.65* | -3.38 | -11.63* | -2.76 | -12.49* |
| Age | 0.01 | 0.79 | 0.01 | **2.07*** | 0.01 | 0.89 | -0.02 | -1.60 | -0.02 | **-2.08*** |
| Sex | 0.40 | 1.96 | 0.64 | **4.90*** | 0.04 | 0.17 | 0.47 | 1.43 | 0.67 | **2.64*** |
| Education | -0.03 | -0.71 | -0.02 | -0.71 | -0.05 | -0.99 | -0.09 | -1.31 | 0.00 | -0.07 |
| Income | -0.24 | **-2.89*** | -0.08 | -1.78 | 0.07 | 0.79 | -0.14 | -1.03 | -0.08 | -0.87 |
| LEC-5 | 0.05 | **5.37*** | 0.04 | **6.33*** | 0.00 | 0.15 | 0.03 | **2.12*** | 0.01 | 0.68 |
| ISS | 0.06 | 1.23 | 0.01 | 0.41 | 0.04 | 0.63 | 0.09 | 1.16 | 0.12 | **2.14*** |
| mCTQ | 0.07 | **7.66*** | 0.05 | **7.37*** | 0.05 | **3.68*** | 0.06 | **4.09*** | 0.05 | **4.10*** |
| CD-RISC | -0.07 | **-6.27*** | -0.05 | **-5.80*** | -0.03 | -1.93 | -0.01 | -0.59 | -0.04 | **-2.89*** |
| PC1 | 5.65 | 1.05 | -1.66 | -0.48 | -10.56 | -1.56 | -8.02 | -0.93 | 8.67 | 1.29 |
| PC2 | -4.68 | -0.90 | -1.28 | -0.41 | 3.19 | 0.61 | 0.10 | 0.01 | 3.38 | 0.57 |
| NDVI | -0.34 | -0.49 | 0.06 | 0.14 | -0.85 | -0.99 | 0.44 | 0.38 | 1.01 | 1.16 |
| ADI | 0.00 | 0.48 | 0.00 | 1.69 | 0.01 | 1.03 | 0.01 | 1.47 | 0.00 | -0.24 |
| AC-PRS | 0.23 | **2.43*** | 0.24 | **3.90*** | 0.09 | 0.78 | 0.37 | **2.42*** | 0.06 | 0.52 |

*Abbreviations:* **AC-PRS:** Ancestry-calibrated PRS; **ADI:** Area Deprivation Index; **CD-RISC***:* Connor-Davidson Resilience Scale; **mCTQ:** modified-Child Trauma Questionnaire; **ISS***:* Injury Severity Score; **NDVI:** Normalized Difference Vegetation Index. **PC***:* top two principal components; *Note:* PC1, PC2, and PRS were z-scored; all other continuous variables were grand mean centered. * $p$ uncorrected < .05.

**eTable 2.** AC-PRS x Ethnoracial Group interaction on PTSD trajectory assignment

| Variable | Trajectory Class (statistical tests relative to the resilient trajectory) | | | | | | | | | |
|---|---|---|---|---|---|---|---|---|---|---|
| | **High Non-Remitting** | | **Moderate Non-Remitting** | | **Delayed** | | **Slow Recovery** | | **Rapid Recovery** | |
| | *Coefficient* | *Wald Z* | *Coefficient* | *Wald Z* | *Coefficient* | *Wald Z* | *Coefficient* | *Wald Z* | *Coefficient* | *Wald Z* |
| (Intercept) | **-2.12** | **-6.52*** | **-0.80** | **-3.68*** | **-2.32** | **-6.99*** | **-3.52** | **-9.39*** | **-2.47** | **-6.59*** |
| Age | 0.01 | 0.80 | **0.01** | **2.03*** | 0.01 | 0.93 | -0.02 | -1.51 | **-0.02** | **-2.05*** |
| Sex | 0.39 | 1.90 | **0.65** | **4.93*** | 0.06 | 0.26 | 0.46 | 1.39 | **0.70** | **2.73*** |
| Education | -0.03 | -0.75 | -0.02 | -0.67 | -0.05 | -1.02 | -0.09 | -1.30 | -0.01 | -0.15 |
| Income | -0.24 | **-2.88*** | -0.09 | -1.84 | 0.07 | 0.81 | -0.13 | -1.02 | -0.08 | -0.85 |
| LEC-5 | 0.05 | **5.34*** | 0.04 | **6.33*** | 0.00 | 0.10 | 0.03 | **2.08*** | 0.01 | 0.62 |
| ISS | 0.06 | 1.19 | 0.01 | 0.42 | 0.03 | 0.54 | 0.09 | 1.21 | **0.12** | **2.09*** |
| mCTQ | **0.07** | **7.62*** | **0.05** | **7.30*** | **0.04** | **3.56*** | **0.06** | **4.12*** | **0.05** | **3.97*** |
| CD-RISC | **-0.07** | **-6.24*** | **-0.04** | **-5.76*** | **-0.03** | **-2.00*** | -0.01 | -0.56 | **-0.04** | **-2.90*** |
| PC1 | 9.82 | 0.93 | 6.12 | 0.83 | -3.19 | -0.33 | **-13.17** | **-9.40*** | 24.91 | 2.29 |
| PC2 | -3.82 | -0.53 | -1.80 | -0.39 | -5.77 | -0.74 | 1.36 | 0.13 | -3.53 | -0.42 |
| NDVI | -0.37 | -0.52 | 0.01 | 0.02 | -0.73 | -0.84 | 0.42 | 0.35 | 1.13 | 1.28 |
| ADI | 0.00 | 0.54 | 0.00 | 1.78 | 0.00 | 0.98 | 0.01 | 1.45 | 0.00 | -0.19 |
| AC-PRS | 0.22 | 1.36 | **0.30** | **3.03*** | 0.02 | 0.14 | 0.47 | 1.84 | 0.29 | 1.46 |
| Hispanic | -0.14 | -0.24 | -0.16 | -0.41 | 0.74 | 1.30 | -0.08 | -0.08 | 0.36 | 0.58 |
| NH Black | -0.22 | -0.47 | -0.35 | -1.09 | -0.29 | -0.61 | 0.29 | 0.77 | -0.69 | -1.37 |
| AC-PRS x Hispanic | -0.17 | -0.47 | 0.14 | 0.60 | 0.02 | 0.07 | 0.00 | -0.01 | -0.67 | -1.86 |
| AC-PRS x NH Black | 0.05 | 0.24 | -0.13 | -1.01 | 0.17 | 0.66 | -0.18 | -0.56 | -0.24 | -0.94 |

*Abbreviations:* **AC-PRS:** Ancestry-calibrated PRS; **ADI:** Area Deprivation Index; **CD-RISC***:* Connor-Davidson Resilience Scale; **mCTQ:** modified-Child Trauma Questionnaire; **ISS***:* Injury Severity Score; **NDVI:** Normalized Difference Vegetation Index; **NH:** Non-Hispanic; **PC***:* top two principal components. *Note:* PC1, PC2, and PRS were z-scored; all other continuous variables were mean centered. *** *p* uncorrected < .05.

**eTable 3.** AC-PRS x NDVI interaction on PTSD trajectory assignment

| Variable | **Trajectory Class** *(statistical tests relative to the resilient trajectory)* | | | | | | | | | |
|---|---|---|---|---|---|---|---|---|---|---|
| | **High Non-Remitting** | | **Moderate Non-Remitting** | | **Delayed** | | **Slow Recovery** | | **Rapid Recovery** | |
| | *Coefficient* | *Wald Z* | *Coefficient* | *Wald Z* | *Coefficient* | *Wald Z* | *Coefficient* | *Wald Z* | *Coefficient* | *Wald Z* |
| (Intercept) | **-2.26** | **-12.89*** | **-1.00** | **-9.27*** | **-2.38** | **-12.64*** | **-3.39** | **-11.63*** | **-2.77** | **-12.49*** |
| Age | 0.01 | 0.79 | **0.01** | **2.07*** | 0.01 | 0.87 | -0.02 | -1.59 | **-0.02** | **-2.08*** |
| Sex | 0.40 | 1.95 | **0.64** | **4.91*** | 0.04 | 0.17 | 0.48 | 1.44 | **0.68** | **2.66*** |
| Education | -0.03 | -0.70 | -0.02 | -0.67 | -0.05 | -1.02 | -0.08 | -1.27 | 0.00 | -0.03 |
| Income | **-0.24** | **-2.89*** | -0.09 | -1.83 | 0.07 | 0.85 | -0.14 | -1.06 | -0.09 | -0.95 |
| LEC-5 | **0.05** | **5.36*** | **0.04** | **6.30*** | 0.00 | 0.19 | **0.03** | **2.10*** | 0.01 | 0.65 |
| ISS | 0.06 | 1.22 | 0.01 | 0.40 | 0.04 | 0.62 | 0.09 | 1.16 | **0.12** | **2.12*** |
| mCTQ | **0.07** | **7.66*** | **0.05** | **7.38*** | **0.05** | **3.67*** | **0.06** | **4.09*** | **0.05** | **4.12*** |
| CD-RISC | **-0.07** | **-6.25*** | **-0.04** | **-5.75*** | **-0.03** | **-1.98*** | -0.01 | -0.57 | **-0.04** | **-2.85*** |
| PC1 | 5.70 | 1.05 | -1.61 | -0.46 | -10.65 | -1.57 | -7.90 | -0.91 | 8.91 | 1.33 |
| PC2 | -4.64 | -0.89 | -1.22 | -0.38 | 3.05 | 0.58 | 0.11 | 0.02 | 3.51 | 0.60 |
| ADI | 0.00 | 0.47 | 0.00 | 1.66 | 0.01 | 1.08 | 0.01 | 1.45 | 0.00 | -0.28 |
| AC-PRS | **0.23** | **2.41*** | **0.24** | **3.95*** | 0.07 | 0.62 | **0.37** | **2.44*** | 0.06 | 0.49 |
| NDVI | -0.31 | -0.44 | 0.08 | 0.17 | -0.90 | -1.04 | 0.33 | 0.28 | 1.15 | 1.30 |
| AC-PRS x NDVI | 0.06 | 0.10 | 0.32 | 0.76 | -0.78 | -0.99 | 0.69 | 0.63 | 0.94 | 1.17 |

*Abbreviations:* **AC-PRS:** Ancestry-calibrated PRS; **ADI:** Area Deprivation Index; **CD-RISC***:* Connor-Davidson Resilience Scale; **mCTQ:** modified-Child Trauma Questionnaire; **ISS***:* Injury Severity Score; **NDVI:** Normalized Difference Vegetation Index; **PC***:* top two principal components. *Note:* PC1, PC2, and PRS were z-scored; all other continuous variables were mean centered. * *p* uncorrected < .05.

### Figures

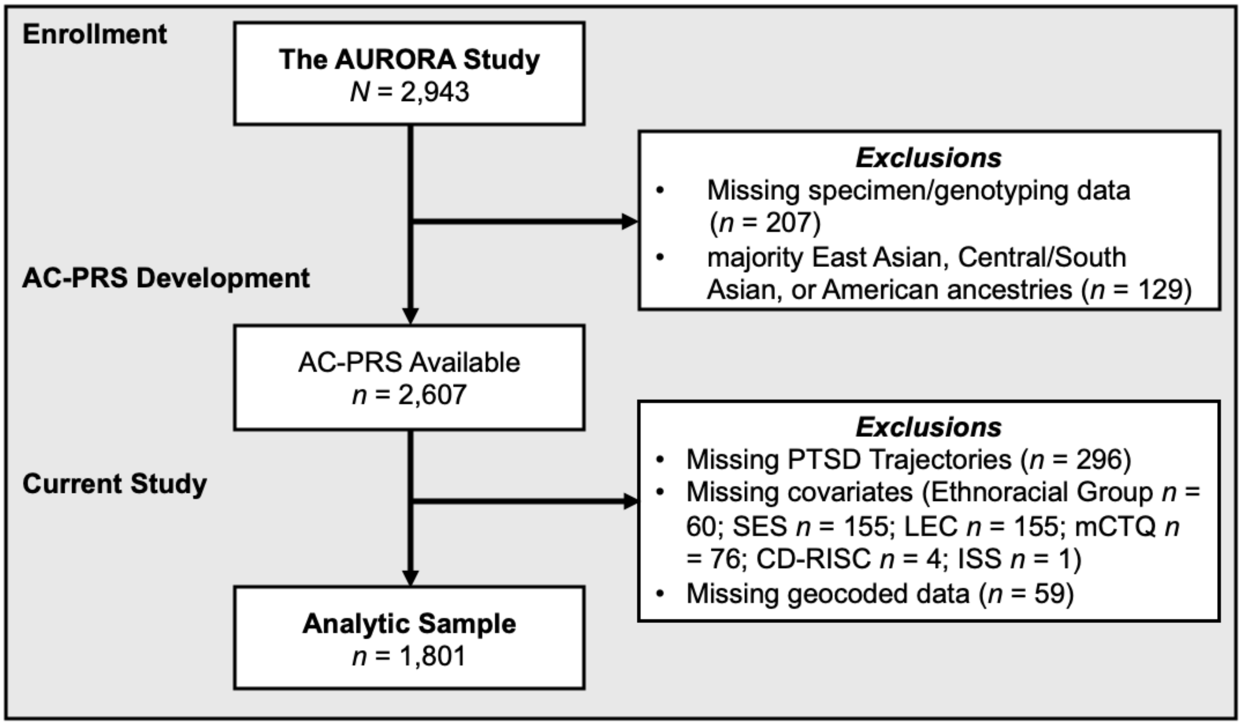


**eFigure 1.** Study flow chart.

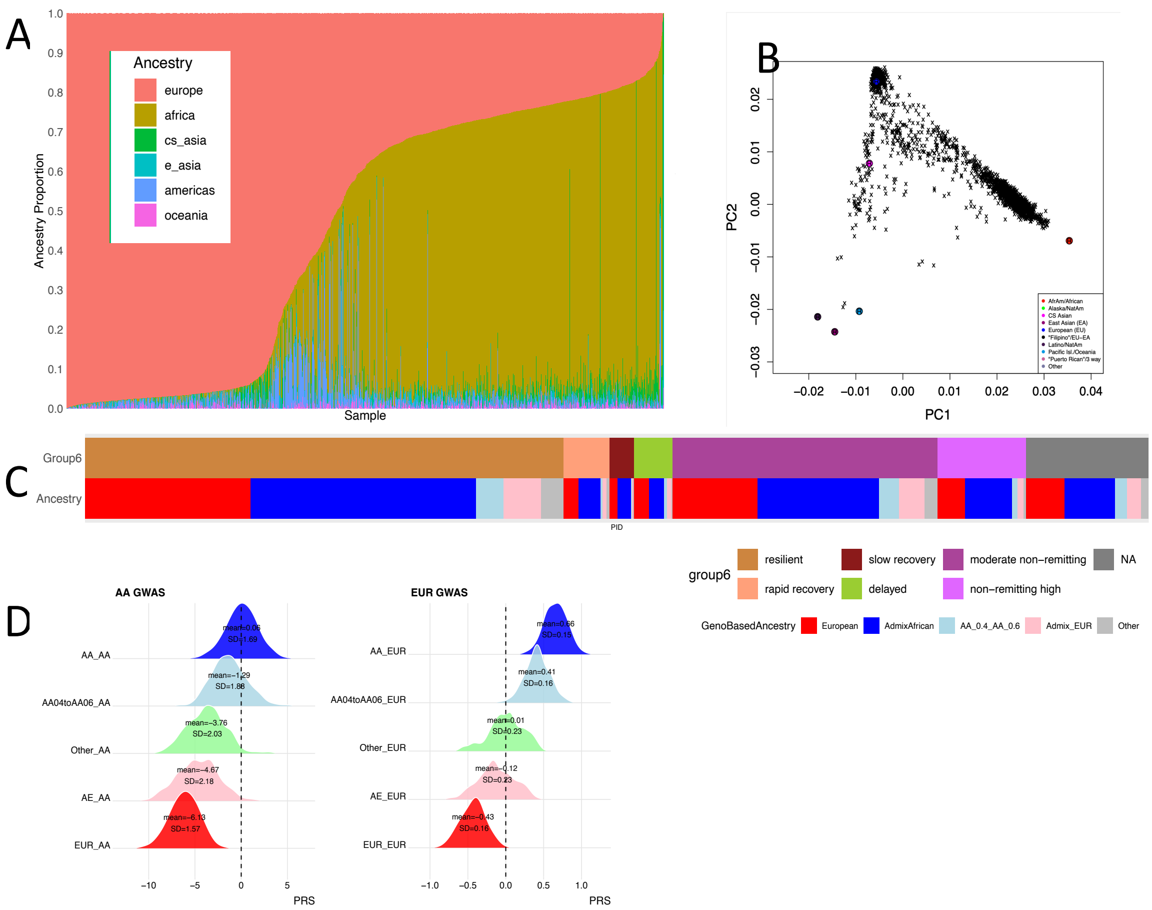


**eFigure 2. Genetic ancestry structure and ancestry-dependent behavior of PTSD PRS in the AURORA cohort. A)** Stacked bar plot displaying the global ancestry proportions (y-axis) of the 2736 subjects (x-axis) from the AURORA cohort. Samples are arranged so that the left side represents subjects of higher European ancestry proportions, while the right side represents participants with higher African ancestry proportions. **B)** Genotype data projected on ancestry marker PCs of the global ancestry pipeline. **C)** Alignment of genetic ancestry assignments with PTSD recovery Group 6 trajectories, highlighting the distribution of ancestry across clinical trajectories. **D)** Distributions of PTSD PRS constructed using ancestry-specific GWAS (*left:* AA GWAS; *right:* EUR GWAS) across ancestry strata, demonstrating substantial ancestry-dependent shifts in PRS mean and standard deviation (SD).

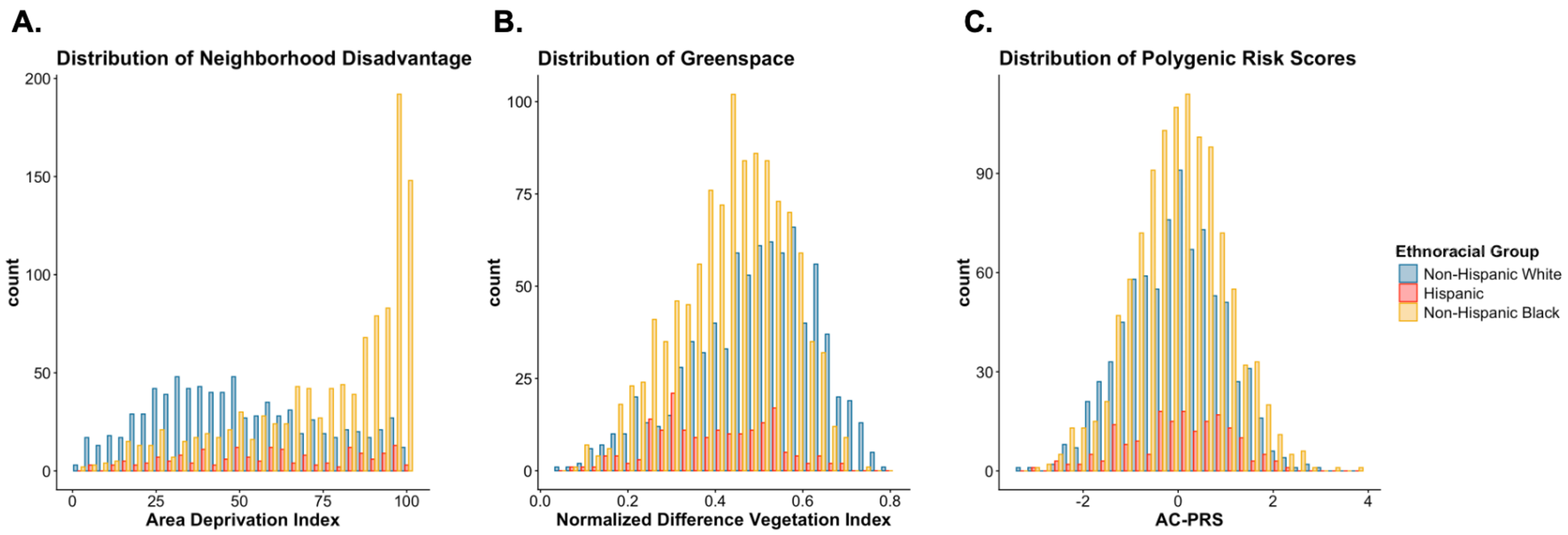


**eFigure 3. Ethnoracial differences in socioenvironmental factors and AC-PRS. [A]** Non-Hispanic White participants lived in significantly more advantaged neighborhoods and were exposed to significantly more residential greenspace **[B]** than non-Hispanic Black and Hispanic participants. **[C]** There were no ethnoracial differences in AC-PRS.

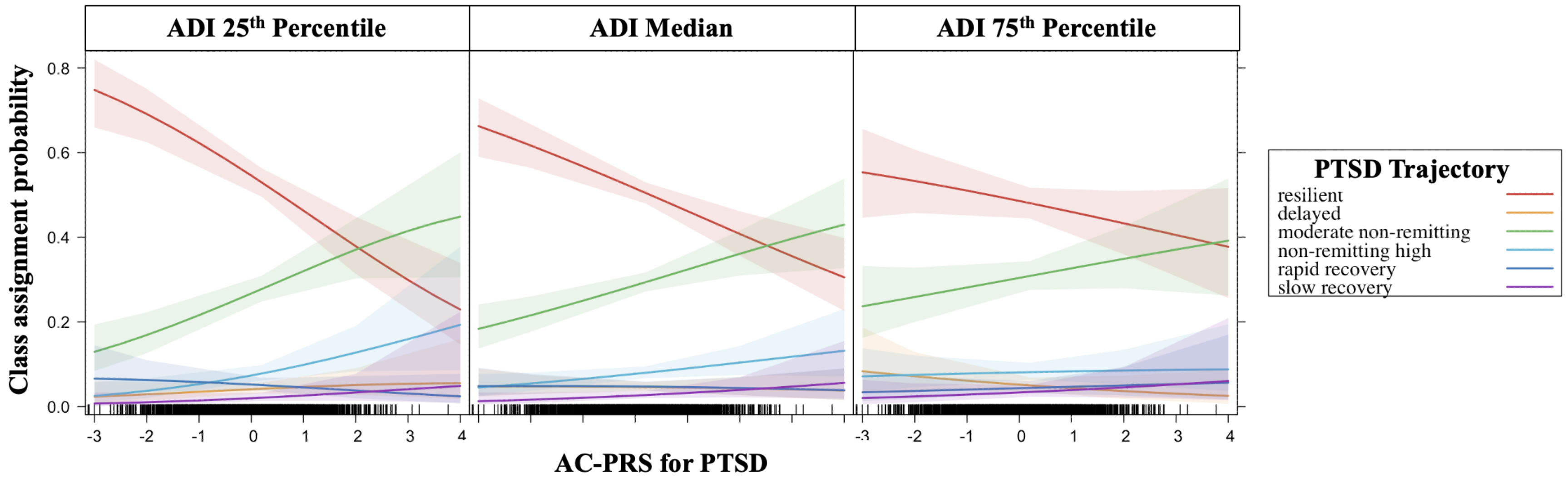


**eFigure 4.** Higher area deprivation index (ADI) attenuates the association between AC-PRS and the likelihood of belonging to the high non-remitting versus resilient PTSD symptom trajectory.
